# THE EFFECT OF DIOXIN ON LOW BIRTH WEIGHT IN SOUTHERN WEST VIRGINIA BETWEEN 1955 – 1969

**DOI:** 10.1101/2020.09.22.20199059

**Authors:** Frank H Annie, Chris K Uejio, Sarah Embrey, Alfred Tager

## Abstract

**Objective:** The effect of low birth weight as a result of dioxin-based exposure has been investigated through the existing literature, but the debate on exact point source and production is still debated. The Monsanto factory located in Nitro, West Virginia produced Agent Orange from 1948 – 1969.

**Methods:** The vital statistics registry from the state of West Virginia of Kanawha county and Putnam county West Virginia. The production of dioxin and 2-4-5T was obtained from the legal files from (Civil Action No. 04-C-465) and the expert witness of Dr. Bruce Bell. In order to understand this relationship a time series analysis was conducted to compare 2-4-5T and dioxin and low-birthweight in Kanawha County and Putnam County, West Virginia.

**Results:** The results suggest dioxin within Kanawha county had a suggestive relationship associated with dioxin production and low birthweight from 1948-1969 had a suggestive relationship of (*p* = 0.042). The comparison county of Putnam had no statistically significant relationship between low birthweight and dioxin production (*p* = 0.203). Agent Orange (2-4-5t) was also considered and measured within the counties of Kanawha and Putnam Counties, within these two counties no statistically significant relationship was observed.

**Conclusions:** In conclusion the association between dioxin and low birthweight still requires more research but this analysis illustrates that there appears to be a suggestive relationship that exists within Kanawha County, West Virginia as related to the production of and effect of the by-product of dioxin.

## Introduction

Preterm birth (i.e., fewer than 37 weeks of gestation) and slower prenatal development produce low-birth-weight infants (1). Newborns weighing 2,500 grams or less are generally classified as low-birth-weight infants and may have slower growth rates, slower cognitive development, and reduced levels of maternal antibodies to fight infections (2,3,4). Low birth weight may also contribute to long-term complications, including chronic heart disease, chronic kidney disease, and a higher risk of developing diabetes later in life (3). An increasing number of low-birth-weight newborns correspondingly increases health care costs for households and health care networks (5,6).

The causes of low birth weight have been determined to be multifactorial and include exposures such as maternal alcohol use, smoking, infection during pregnancy, drug abuse, maternal age less than 17 years or greater than 35 years, and race (7,8, 9, 10). The mother’s health status can also play a significant role in the development process, as any related illnesses can alter the development process of the fetus (11). Other external variables that have been examined include toxic chemical exposures such as dioxins, lead, mercury, and PCBs (11).

Current literature indicates a possible link between low birth weight in newborns and dioxin exposure (12, 13, 14). Dioxins are a class of chemical by-products generated in various manufacturing processes, such as the production of rubbers and synthetic plastics. At the Monsanto plant in Nitro, West Virginia, which is the focus of this study, dioxins were generated during the production of sodium 2,4,5-Trichlorophenate (NaTCP) (15,16). The estimated amount of dioxin produced from 1948 to 1969 was approximately 0.47 parts per million (17). Depending on the subclass, dioxin exposures below 0. 1–1.0 parts per trillion may not cause adverse health effects (18).

The primary avenue of human exposure is through consuming contaminated foods (19); however, in populations with consistent dioxin exposure, dermal absorption may also be an important exposure route. The pathophysiology of how the body processes dioxin differs from person to person, but the substance is metabolized and broken down within the liver and absorbed into fatty cells. The rate of breakdown depends on the subclass of dioxin and the amount of fat within the body (20). Dioxin’s chemical structure is similar to some hormones and may bind to cellular aryl hydrocarbon receptors and immediately affect human health (19). Dioxin can remain in the tissue for many years depending on fat content (20). Exposure at sufficient levels may cause health problems such as cancer, male reproduction toxicity, female reproduction toxicity, effects on unborn fetuses, skin disorders, and other metabolic and hormonal abnormalities (19,21). This study builds upon previous work where areas that produced or have high dioxin concentrations also report elevated rates of low birth weight compared to neighboring counties (22, 6). Some studies suggested that excessive exposure over a short period may also negatively impact birth outcomes (23).

This study focuses on a point dioxin exposure source in a United States community with a lower dioxin production (of approximately 0.47 parts per million) than Vietnam. During the Vietnam War, larger dioxin exposures caused adverse health outcomes these exposure quantities could range from 0.001 to 1.0 parts per million depending on the level of contamination in the region (19). This study investigates the relationship between Kanawha County’s and neighboring Putnam County’s low birth weight rates and the Monsanto plant dioxin production from 1955 to 1969. Other dioxin studies focus primarily on indirect dioxin and PCB exposures from consuming food (e.g., fish) and low newborn birth weight (24). This study also examines the synchronous relationship between dioxin production and low birth weight whereas other studies examine the lingering health effects of existing dioxin contamination. Furthermore, this study separately compares dioxin production against the proportion of low-birth-weight children while controlling for independent risk factors such as median household income.

## Methods

### Study Area

The study area includes the residents of Kanawha County and Putnam County, West Virginia. Kanawha County consists of Nitro, Charleston (the state capitol), and over 100 towns and small cities located near the Monsanto plant (see Figure 1). From 1948 to 1969, the U.S. government commissioned chemical companies such as Monsanto to produce Agent Orange in Nitro, West Virginia. By 1969, the U.S. government had discontinued using the substance for any large-scale military operations in Vietnam, and Monsanto subsequently halted Agent Orange production at its Nitro plant. To provide a comparison, the study tests whether Kanawha County dioxin production is related to low birth weight in Putnam County. This county was compared because of its geographical proximity to Kanawha County as well as the low birthweight data was also available from Putnam County as well. No additional point sources of pollution were in the Putnam County area (see Figure 1).

**Figure 1.**
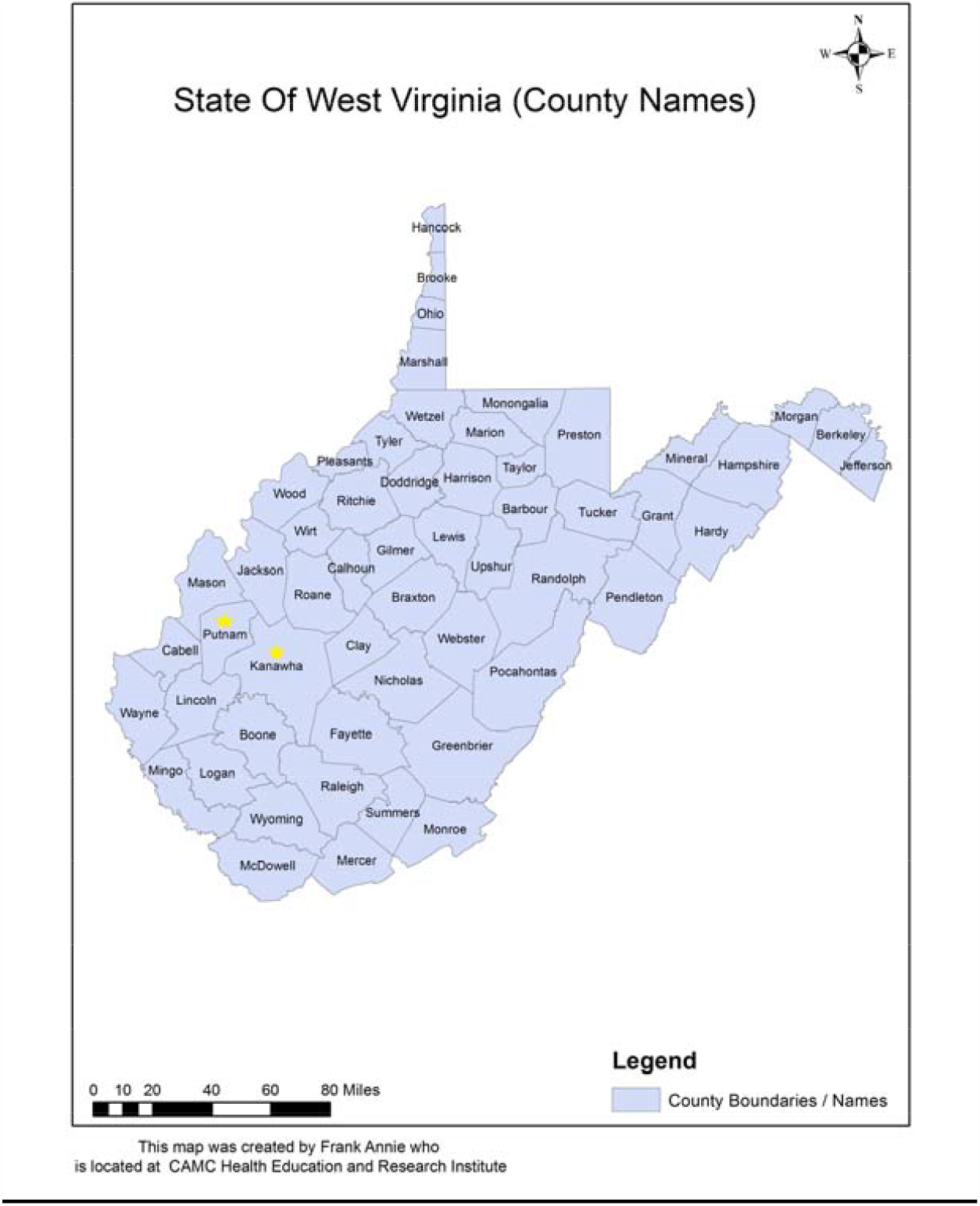
Kanawha County, West Virginia, which includes the county seat of Charleston, West Virginia (the state capitol), had a census of 252,925 in 1960. The area is defined by a massive chemical industry as well as the coal industry. Kanawha County also includes large banks, hospitals, and universities. Putnam County, which includes the county seat of Winfield and the largest city of Hurricane, had a census of 23,561 in 1960. Putnam County included several towns and later became a suburban area between Kanawha County and Cabell County, West Virginia.

### Data

This study’s primary data sources include birth records from the State of West Virginia Vital Statistics Registry and a court document from the Putnam County Court of West Virginia, the former of which provided the annual total number of births and low-birth-weight infants along with the proportion of low-birth-weight infants in the study counties. The study involved subjects born between 1955 and 1969 who were issued a new birth certificate in Kanawha or Putnam County. The Vital Statistics Registry did not compile a database of birth records before 1955. Moreover, due to a structure fire, additional documents from 1970 to 1985 were destroyed or damaged and could not be obtained as they no longer existed. The registry did not include those who resided in the county but those who went to a medical facility outside the county to give birth.

The second data source is the Putnam County Court document against the Agricultural Division of Monsanto. From 1948 through 1969, this report estimated dioxin production loss to the environment through burning, sewer loss, production, and waste disposal, which was estimated to be 10,000 pounds. Dr. Bruce Bell, an expert witness for the plaintiff, reported the estimates during the court case discovery period for the plaintiff (Civil Action No. 04-C-465). Dr. Bell’s estimates were generated from the Monsanto plant blueprints and sales receipts. The Calwell Practice PLLC, Law and Arts Center West in Charleston, West Virginia, commissioned the report.

This study also considered statewide annual median household income provided by the United States Census Bureau. Median household income was adjusted and standardized for inflation from 1955 to 1969. Income may be a surrogate marker for access to health care, health literacy, smoking, and alcohol usage, all of which may influence low birth weight (25). In fact, the correlation between household income and low birth weight has been established within the literature (26). The study could not control for annual rates of smoking and alcohol usage, as these were unavailable for the study period and region.

### Statistical Analysis

For each county, we created separate multiple variable time series models to relate dioxin production to low birth weight while controlling for median household income. Epidemiology commonly uses time series analysis to describe a short-term exposure to a health outcome (27). The technique compares rates in the same study population over time, which naturally controls for slowly changing low birth weight confounders.

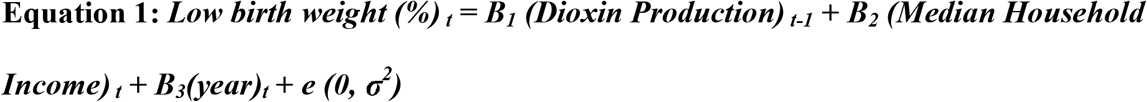

In Equation 3, low birth weight is the percentage of low-birth-weight infants, and the exposure variable is dioxin production. The equation also controls for median household income and long-term temporal trends (study year). To further account for the gestational lag between exposure and birth, each exposure variable was lagged one year. For example, exposure in 1955 would influence birth weight in 1956; the subscript *t* thus refers to the year sequence beginning at 1956, or 1955 for *t-1*. The time series model assumptions were then verified with standard diagnostic tests. A histogram confirmed that the residuals were normally distributed. Furthermore, autocorrelation and partial autocorrelation functions verified that the time series did not contain significant residual temporal autocorrelation. Finally, Akaike and Bayesian Information Criterion measured the time series model fit.

## Results

Tables 1 and 2 report the regression results from the best fitting time series models. Both models fulfilled the time series model assumptions. The Kanawha County regression model tested the association between annual dioxin production and the percentage of low-birth-weight infants (see Table 1). Based on the beta coefficient, every 10 pounds of dioxin produced an increased incidence of low birth weight by 0.41%. Moreover, each $10,000 USD increase in household income decreased the incidence of low birth weight by 4.5% (*p* = 0.005). Figure 2 plots the relationship between observed and time series–fitted low birth weight for dioxin production.

**Table 1.** Results of the time series analysis on dioxin production and low birth weight in Kanawha County, West Virginia.

| Variable | Coefficient | Standard<br>Error | T | P > t | (95%) Confidence<br>Interval |
| --- | --- | --- | --- | --- | --- |
| Dioxin Production | 0.0411 | 0.0178 | 2.31 | 0.042 | 0.0018–0.0804 |
| Median Income | –4.528 | 1.1938 | –3.48 | 0.005 | –6.7805––1.5251 |
| Year | 0.2457 | 0.122 | 2.19 | 0.051 | –0.0012–0.4928 |

**Table 2.** Results of the time series analysis on dioxin production and low birth weight in Putnam County, West Virginia.

| Variable | Coefficient | Standard<br>Error | T | P > t | (95%) Confidence<br>Interval |
| --- | --- | --- | --- | --- | --- |
| Dioxin Production | 0.0053 | 0.0039 | 1.35 | 0.203 | –0.0033–0.0140 |
| Median Income | –0.0001 | 0.0003 | –0.83 | 0.424 | –0.0001–0.0004 |
| Year | 0.1827 | 0.2767 | 0.66 | 0.522 | –0.4203–0.7857 |

**Figure 2.**
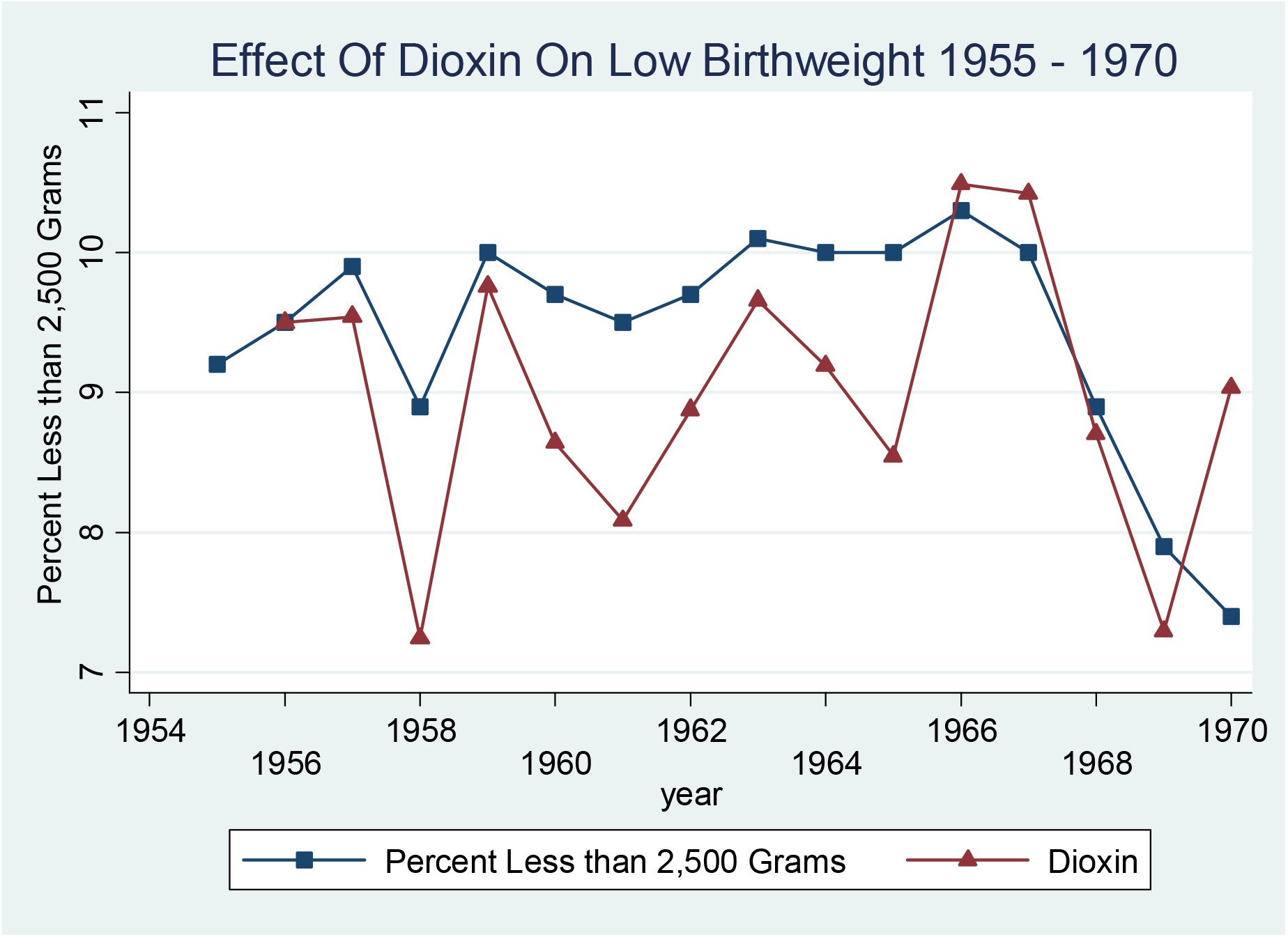
Time series analysis on low birth weight compared to dioxin production in Kanawha County, West Virginia, from 1955 to 1969 (*P* = 0.042).

Table 1 reports the regression results of the best fitting time series model of Putnam County. Dissimilar to Kanawha County’s regression table, this one indicates no significant results associated with the acute exposure of dioxin and low birth weight. Figure 3 plots the relationship in the county between the observed acute dioxin exposure and the time series–fitted low birth weight time series analysis.

**Figure 3.**
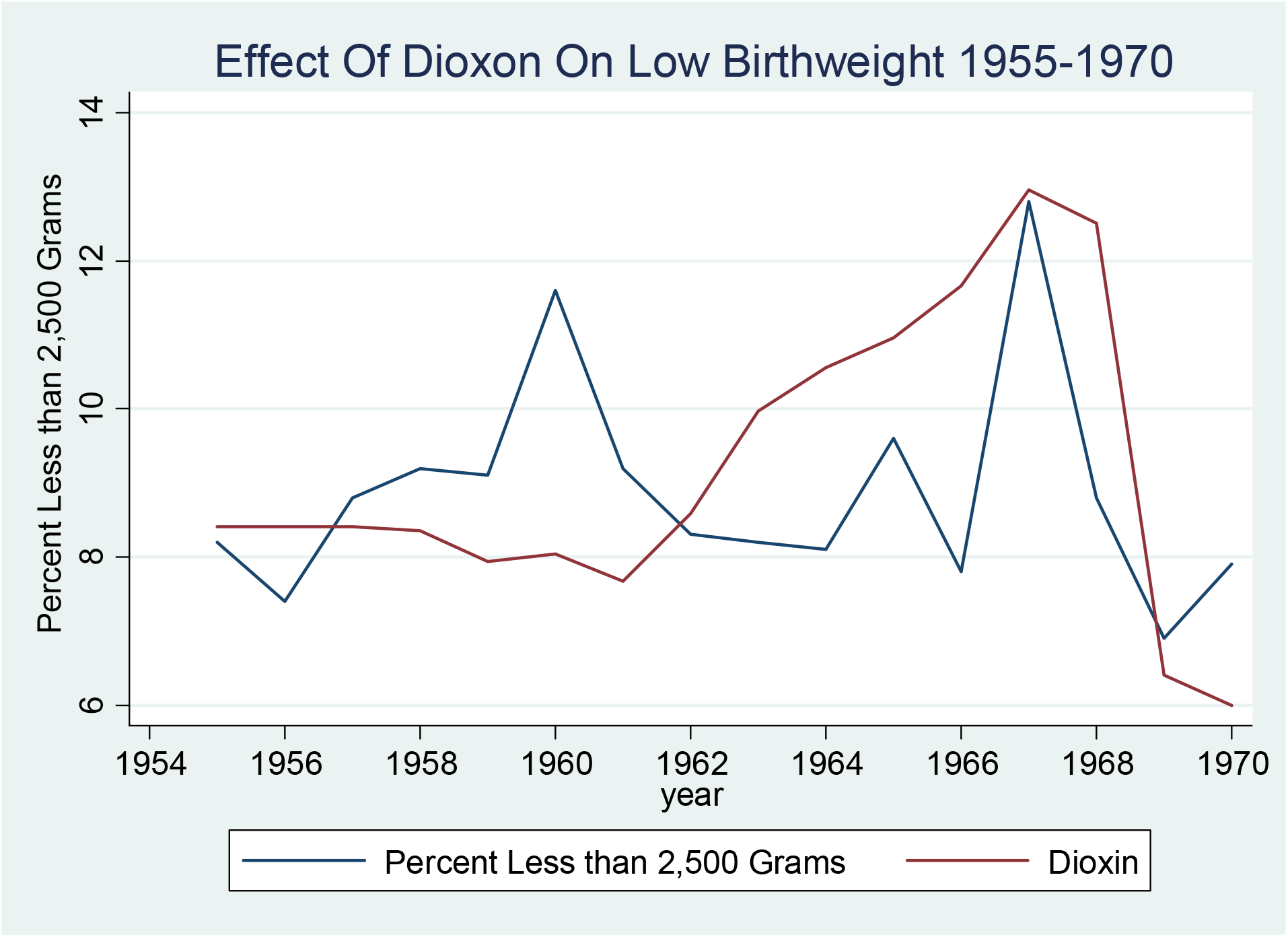
Time series analysis on low birth weight compared to dioxin production in Putnam County from 1955 to 1969 (*P* = 0.203).

## Discussion

This study found a suggestive relationship between an increase of dioxin from the production of Agent Orange (2,4,5-T) (see Appendix) and an acute increase of low birth weight in Kanawha County, West Virginia (*P* = 0.042). Individuals with lower socioeconomic status consistently tend to have infants with lower birth weights (25,28, 29, 30, 31,). A comparison county, Putnam County, located west of Kanawha County, was analyzed during the same period. Results found that there was no association between low birth weight and the production of dioxins (*P* = 0.203) in Putnam County. This outcome provides additional evidence that Kanawha County dioxin exposure is not a proxy for an unmeasured time-varying exposure.

Our study results suggest that dioxin production may increase the incidence of low birth weight over time. Again, this study builds upon previous work where areas that produced or have high dioxin concentrations also report elevated rates of low birth weight compared to neighboring counties (32,33). These previous two studies examined exposure related to the consumption of food or working in dioxin-related industries. For instance, the Rylander 1995 study assessed the high use of contaminated food from the (Sweden) Baltic Sea that was associated with an increased risk of low birth weight in the community; the Rylander 2000 study further supported the link between the intake of dioxin from fish and an increased risk of low birth weight. However, neither study estimated a range of dioxin exposure levels that corresponded to the risk of low birth weight infants. Further studies such as the Tsikimori 1968– 2004 work explored the link between exposure to rice contaminated with Yusho oil and preterm births and induced abortions in Japan, researching the period from 1968 to 2004, when dioxin exposure had ceased. The study found that women exposed to dioxins had a twofold increase in preterm pregnancy and an increase in induced abortions five times greater than the baseline number.

In the study area and period, dioxin exposures were estimated at 0.47 parts per million (Civil Action No. 04-C-465). This level of exposure is generally similar to that found in international studies of dioxin exposure and adverse newborn outcomes. Karmaus’s 1995 study, conducted in Germany, more precisely measured dioxin exposures and low birth weight: newborns with median concentrations of dioxin in blood samples of 0.5 parts per million demonstrated an average decrease of 175 grams and 2 centimeters from their birth measurements compared to nonexposed infants. Relatedly, studies suggest acute dioxin exposures of approximately 0.3 parts per million increase the risk of urinary tract birth defects in France (32).

Our studies are partially consistent with studies on laboratory animals, specifically rat studies illustrating reduced size of offspring (34,35). Dioxin binds to the fetus’s cellular aryl hydrocarbon receptor (AhR), which activates a gene that influences growth hormones. Dioxins can act at the AhR due to their structural similarity to other endogenous hormones in the human body. This occurrence in turn causes random growth variations that lead to the reduced formation and growth of an infant (36). Research on AhR receptors confirms a correlation between dioxin-linked gene modification and chronic effects such as a compromised immune system and soft tissue damage (37).

The primary mechanism of dioxin exposure is consuming food contaminated with fine dioxin and other chemical powders that circulate in the area. The dioxin was released into the water and air of the surrounding community through waste disposal and incineration practices (38, 39). The fine dioxin powder adheres to clothing and can accumulate inside buildings and cause adverse health outcomes over time or have a more immediate effect (40, 41).

This section describes the limitations of the population-based study, which used existing secondary data sources to investigate the relationship between dioxin production and low birth weight. The study also used a time series analysis to account for as many limitations as possible. These models control for other low birth weight risk factors that change slowly over time; however, the study design cannot control for the effects of other chemicals in the study area with similar production schedules to dioxins. For example, the DuPont factory in Dunbar, West Virginia, was five miles upstream from the Monsanto site, and data regarding the production of substances at this plant could not be obtained for this study.

Other noteworthy issues concern births that occurred at institutions outside Kanawha County and the time when birth certificates were issued. For instance, it should be noted that the study period covers the time before the interstate highway system was constructed. It therefore may have been difficult to reach the next closest hospital outside the patient’s home county, and for this reason, relatively few patients would have been misclassified into the incorrect county. A time lag may have also occurred between home birth and birth registration, as only children born in a hospital would have been issued a birth certificate immediately. Conducting the time series analysis at yearly intervals would minimize—but not eliminate—birth timing misclassification error.

Our study relied on testimony from a single source to estimate chemical production and used median household income as a surrogate for health status and behaviors such as alcohol and tobacco usage. Controlling for these risk factors would strengthen the study results, but historical local alcohol and tobacco use rates were not systematically collected. We examined national alcohol and tobacco use trends over time to test whether they exhibit a similar pattern to dioxin production. Smoking prevalence was increasing during the mid-1950s, before the study period; during the study period, it remained relatively constant until the mid-1970s, with a gradual downward trend when bans on smoking commercials were introduced and smoking cessation began to be promoted (42). A National Institute of Alcohol Abuse and Alcoholism study illustrates that alcohol consumption increased across the study period (42). Increased alcohol consumption could lead to increased low birth weight depending on levels of consumption (43). If regional statistics mirrored national alcohol and tobacco consumption trends, these risk factors did not mimic the dioxin production schedules.

## Conclusion

This study illustrates that a higher exposure level of 0.47 parts per million can influence low birth weight within the surrounding area. Furthermore, the findings of this study reveal a suggestive relationship (*p* = 0.042) as compared to low birth weight within Kanawha County, West Virginia. A comparison county, Putnam County, was included to provide further evidence of an association between dioxin exposure and low birth weight, but this test provided no statistical significance (*P* = 0.203). Dioxins and dioxin-like substances are prevalent within our environment even now, and the findings of this study suggest that there might be an effect. Further research is needed to understand the continued effect within the environment, specifically in the Appalachian Region. Based on existing literature, dioxin is still within the environment of Kanawha County, West Virginia; at high levels and with a multidecade half-life, it will remain in the environment for years to come.

## Data Availability

Data can be obtained upon request

### Appendix

**Table A1.** Results of the time series analysis of 2,4,5-T production and low birth weight.

| Variable | Coefficient | Standard Error | T | P > t | (95%) Confidence Interval |
| --- | --- | --- | --- | --- | --- |
| 2,4,5-T Production | 0.0019 | 0.0009 | 1.95 | 0.077 | −0.0002–0.0041 |
| Median Income | −4.0929 | 1.2525 | −3.27 | 0.007 | −6.8497–−1.3362 |
| Year | 0.2492 | 0.1178 | 2.12 | 0.058 | −0.0100–0.5086 |

There is a positive but marginally significant relationship between low birth weight and 2,4,5-T production (*p* = 0.077) (see Table A1): Based on the beta coefficient, every 10,000 pounds of 2,4,5-T produced increased the incidence of low birth weight by 0.19%. Similar to the dioxin model, higher median household income significantly decreased the proportion of low-birth-weight children (*p* = 0.007). For every $10,000 increase in median household income, low birth weight incidence decreased by 4.0%. Figure A1 plots the relationship between observed and time series–fitted low birth weight for 2,4,5-T production. The model demonstrates a supportive relationship, but certain years (e.g., 1968 and 1969) do not exhibit a strong relationship between production and low birth weight.

---

**Figure A1.**
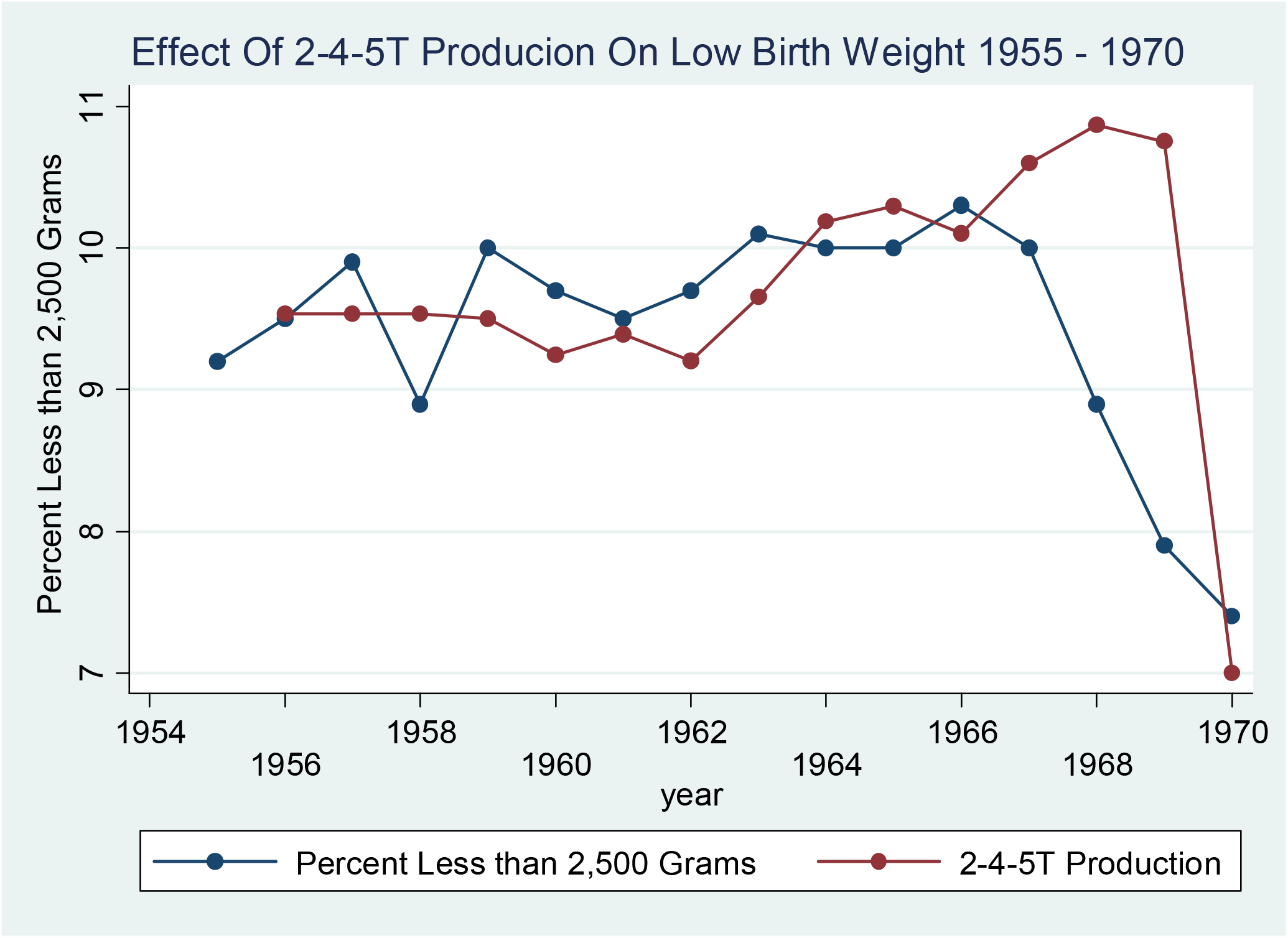
Time series analysis of low birth weight 2,4,5-T production.

